# Language-Related Differences in Prenatal Depression Screening Uptake, US Midwest 2019-2024

**DOI:** 10.64898/2026.04.07.26350332

**Authors:** Amanda Luff, Anne Rivelli, Akaninyene Noah, Emily Malloy, Ravi Mishra, Veronica Fitzpatrick

## Abstract

Prenatal depression is a substantial contributor to maternal morbidity, and screening is an entry point to psychiatric assessment and treatment during pregnancy. Following updated guidelines and quality metrics for prenatal depression screening, we evaluated whether screening uptake differed by preferred language within a large U.S. healthcare system. We used electronic health record data to identify a retrospective cohort of deliveries at or beyond 20 weeks gestation in 2019-2024. We used logistic regression with a language-year interaction to estimate the adjusted marginal probabilities of screening by language preference. Among 99,526 pregnancies (82,632 individuals), screening increased substantially over time but increases differed across language groups (p<0.001). In 2019, screening probabilities were similar (English 0.50; Spanish 0.48; Another Language 0.50). By 2024, probabilities diverged (English 0.81; Spanish 0.66; Another Language 0.71). Unequal screening uptake can systematically under-identify prenatal depression among patients with non-English language preference, with implications for equitable access to psychiatric care.

## Introduction

Prenatal depression impacts nearly 2 in 5 pregnancies in the United States and when untreated is associated with reduced engagement in prenatal care, preterm birth, postpartum depression, and impaired child neurodevelopmental outcomes.^1–3^ Perinatal depression is a leading contributor of maternal mortality, with death by suicide accounting for up to 20% of postpartum deaths.^4,5^ Because most mental health symptoms during pregnancy are first identified in obstetric settings, prenatal depression screening is an important pathway to further assessment, follow-up, and referral for psychiatric evaluation and treatment.^6,7^ The American College of Obstetricians and Gynecologists issued its first recommendation for universal prenatal depression screening in 2015 and reaffirmed this in 2018.^8,9^ In 2019 the National Committee for Quality Assurance added Prenatal Depression Screening and Follow-up as a quality measure.^10^ As systems expand screening through standardized workflows and performance measurement, equitable delivery is essential to avoid systematic under-identification of prenatal depression in groups facing barriers to care. Language is a known barrier to prenatal depression screening.^11^ This study evaluated the relationship between language preference and uptake of prenatal depression screening within a large, diverse U.S. healthcare system.

## Method

This retrospective cohort comprised pregnancies resulting in deliveries at or beyond 20 weeks gestation between 2019 and 2024. The study was conducted within a large, vertically integrated healthcare system in the U.S. Midwest comprising 19 delivery facilities. We included pregnancies that initiated prenatal care in the first or second trimester. All data were extracted from the electronic health record (EHR). We excluded pregnancies with missing variables of interest using listwise deletion (<5% of the sample). We reported findings following the Strengthening the Reporting of Observational Studies in Epidemiology (STROBE) guidelines.^12^

Prenatal depression screening was determined by presence of a depression screening instrument in the EHR during the PNC period. We used logistic regression with cluster-robust standard errors to account for individuals contributing multiple pregnancies.

Using an interaction term, we evaluated whether the association between language preference and receipt of prenatal depression screening varied by year of delivery, adjusting for maternal age, race/ethnicity, insurance type, parity, gestational age, singleton versus multiple gestation, and the state in which PNC occurred. We calculated and graphed marginal probabilities of receiving prenatal depression screening across years for each language group.

The authors assert that all procedures contributing to this work comply with the ethical standards of the relevant national and institutional committees on human experimentation and with the Helsinki Declaration of 1975, as revised in 2013. All procedures involving human subjects were approved by The Wake Forest School of Medicine Institutional Review Board (IRB00129720) with a waiver of informed consent due to retrospective methods.

## Results

Between 2019 and 2024, 99,526 eligible pregnancies among 82,632 individuals resulted in a delivery in this healthcare system. An additional 846 pregnancies (0.84%) were excluded because of missing variables used in the analysis. Among the included pregnancies, 37% received prenatal care in Illinois and 63% in Wisconsin. Pregnant people reported 76 preferred languages; because most languages had small counts, we report language preference as English (94%), Spanish (3%), and Another Language (3%) to ensure stable estimation.

In the model estimating odds of receiving prenatal depression screening, the interaction between language and year was statistically significant (p<0.001). In 2019, marginal probabilities of prenatal depression screening were similar across groups: English, 0.50 (95% CI, 0.49–0.51); Spanish, 0.48 (95% CI, 0.41–0.56); and Another Language, 0.50 (95% CI, 0.42–0.58). These probabilities diverged over time (**Figure 1**). By 2024, the probability of screening increased to 0.81 (95% CI, 0.80–0.81) for English-language preference, 0.66 (95% CI, 0.63–0.70) for Spanish-language preference, and 0.71 (95% CI, 0.68–0.74) for another language preference.

**Figure 1.**
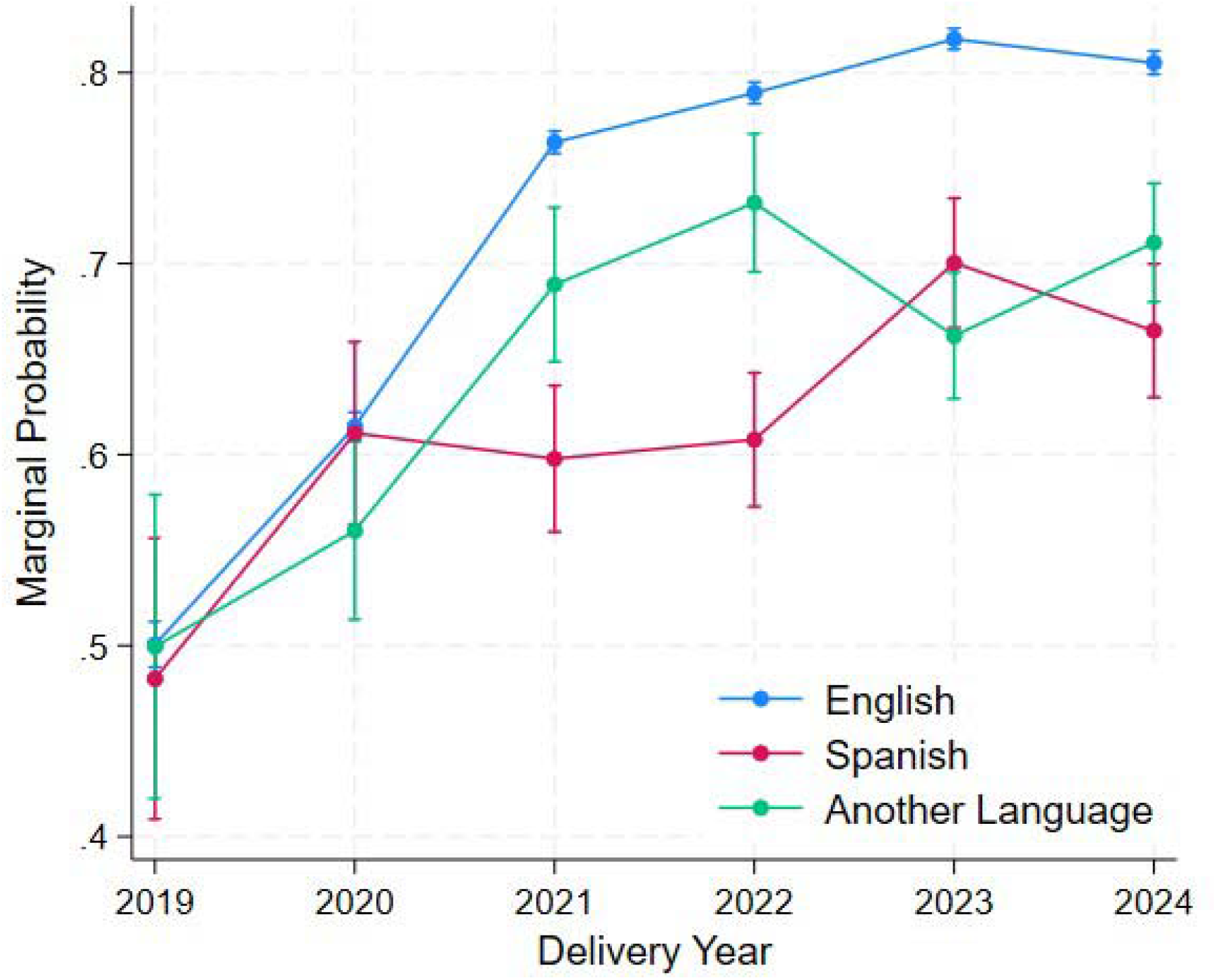
Marginal Probabilities of Prenatal Depression Screening by Preferred Language and Delivery Year, 2019-2024.

## Discussion

Prenatal depression screening increased from 2019 to 2024, but improvements were not equal across language groups, resulting in larger screening gaps by 2024 for individuals with non-English language preference. Screening probabilities were similar across language groups early in the evaluation period, indicating that disparities emerged as screening expanded rather than reflecting persistent baseline differences. Prenatal depression screening is typically conducted in obstetric care as a primary pathway through which perinatal depression is identified and patients are connected to mental health services.^6^ From a health-systems perspective, screening gaps represent missed opportunities for earlier intervention, including initiation of evidence-based psychotherapy and/or pharmacotherapy, care management, and collaborative care approaches. If screening is less consistently completed for people with non-English language preference, fewer patients may be identified during routine prenatal care, potentially leading to patients with non-English language preference presenting with higher-acuity mental health conditions.

That disparities emerged as screening expanded suggests that implementation improved overall completion without fully addressing language access. Incorporation of screening into performance metrics may increase uptake, but without routine stratification by language preference, aggregate reporting can obscure widening inequities. Screening programs often use EHR prompts, standardized workflows, and embedded instruments, which may be built first for English-preference patients. When interpreter support or translated tools are needed, screening can require additional steps, such as locating validated translations and different documentation processes, increasing burden for patients and clinical teams. The presence of many preferred languages with small counts highlights a structural challenge to equitable screening, as validated instruments may be unavailable for some language groups. In addition, small numbers across many languages can limit routine health system attention to any single group, even when the full patient population with non-English language preference is substantial.

Strengths of this study include the large cohort across multiple years within a diverse health system and use of EHR-derived measures to evaluate real-world screening implementation. The analytic approach accounted for individuals contributing multiple pregnancies and examined whether the association between language preference and screening changed over time, which directly addresses how inequities may emerge during program expansion. A limitation to this research is that screening was identified based on documentation of a screening instrument in the EHR; this may not fully capture whether screening was completed as intended, understood by the patient, or administered with interpreter support when needed. Second, language preference recorded in the EHR may not perfectly reflect language proficiency, literacy, or degree of need for language support services, and the “Another Language” category necessarily aggregates heterogeneous language groups. Third, this analysis evaluated receipt of screening rather than downstream steps such as severity of symptoms, follow-up, referral completion, or treatment initiation; therefore, we cannot directly quantify how screening gaps translated into differences in mental health service access or outcomes. Finally, results reflect a single health system and may not generalize to settings with different screening workflows or language-access resources.

In summary, prenatal depression screening increased substantially from 2019 to 2024, yet language-related inequities widened as screening expanded. Given that mental health–related morbidity is a substantial contributor to overall maternal morbidity and that screening is a key entry point to mental health services, equitable screening implementation is a health-systems and psychiatric care priority.^3,5^ Stratifying quality metrics by language preference and designing multilingual screening workflows may help ensure that screening expansion translates into equitable access to essential perinatal mental health care.

## Data Availability

The data that support the findings of this study are available on request from the corresponding author, subject to institutional data sharing procedures. The data are not publicly available to protect patient privacy.

## CRediT Statement

**Amanda Luff**: Conceptualization, Formal analysis, Writing - Original Draft; **Anne Rivelli**: Conceptualization, Funding acquisition, Supervision, Writing - Original Draft; **Akaninyene Noah**: Data Curation, Writing - Review & Editing; **Emily Malloy**: Conceptualization, Writing - Review & Editing; **Ravi Mishra**: Conceptualization, Writing - Review & Editing; **Veronica Fitzpatrick**: Supervision, Writing - Original Draft

